# An assessment of census-tract level socioeconomic position as a modifier of the relationship between PM 2.5 concentrations and cardiovascular emergency department visits in Missouri

**DOI:** 10.1101/2023.09.25.23296034

**Authors:** Zachary H. McCann, Howard H. Chang, Rohan D’Souza, Noah Scovronick, Stefanie Ebelt

## Abstract

Ambient PM2.5 exposure elevates the risk for cardiovascular disease morbidity (CVDM). The aim of this study is to characterize which area-level measures of socioeconomic position (SEP) modify the relationship between PM2.5 exposure and CVDM in Missouri at the census-tract (CT) level. We use individual level Missouri emergency department (ED) admissions data (n = 3,284,956), modeled PM2.5 data, and yearly census tract data from 2012-2016 to conduct a two-stage analysis. Stage one uses a case-crossover approach with conditional logistic regression to establish the baseline risk of ED visits associated with interquartile range (IQR) changes in PM2.5. In the second stage, we use multivariate meta-regression to examine how census tract level SEP modifies the relationship between ambient PM2.5 exposure and CVDM. We find that overall, ambient PM2.5 exposure is associated with increased risk for CVDM. We test effect modification in statewide and urban census tracts, and in the warm-season only. Effect modification results suggest that among SEP measures, poverty is most consistently associated with increased risk for CVDM. For example, across Missouri the highest poverty CTs are at an elevated risk for CVDM [OR = 1.010 (95% CI 1.007, 1.014)] compared to the lowest poverty CTs [OR = 1.004 (95% CI 1.000, 1.008)]. Other SEP modifiers generally display an inconsistent or null effect. Overall, we find some evidence that area-level SEP modifies the relationship between ambient PM2.5 exposure and CVDM, and suggest that the relationship between air-pollution, area-level SEP, and CVDM may be sensitive to spatial scale.

---

Exposure to ambient fine particulate matter <2.5 μm in aerodynamic diameter (PM_2.5_) is associated with increased levels of cardiovascular disease morbidity (CVDM). [1–3] Identifying the populations at increased risk of exposure to health effects from PM_2.5_ is an important step in public health protection from this key and complex air pollutant.

The Environmental Protection Agency (EPA) has identified several characteristics of area-level socioeconomic position (SEP) that put communities at risk for elevated exposure to environmental contaminants, including ambient PM_2.5_. These characteristics include high concentrations of people of color (POC), individuals with low incomes, unemployment, households with limited English proficiency, low educational attainment, and elderly residents. [4] Recent research confirms that several EPA area-level SEP indicators, including areas with larger proportions of POC, highly concentrated poverty, and high unemployment rates are associated with increased PM_2.5_ exposure. [5–9] Despite clear evidence of the deleterious cardiovascular effects of ambient PM_2.5_, and an unequal burden of PM_2.5_ exposure across social strata, evidence surrounding the role of area-level SEP in modifying the relationship between PM_2.5_ exposure and CVDM is sparse with mixed findings. [10–13]

The use of Zone Improvement Plan (ZIP) codes and ZIP Code Tabulation Areas (ZCTAs), the most commonly available spatial units for these types of studies, may be one reason for inconsistent results. [14] Population-based short-term health effect studies are conducted by leveraging large administrative databases (e.g., hospital billing data), where ZIPs or ZCTAs are often the finest spatial resolution available for patient residential information.

Emerging evidence suggests that census tracts (CTs) may be better suited for detecting how area-level SEP modifies air pollution-related CVDM risk. [1,15–17] Compared to ZIPs and ZCTAs, CTs contain relatively homogenous populations with respect to income, housing, and racial characteristics, map well to other geographic features (e.g., counties, states, census blocks), and are better able to capture population-level statistical data. [18,19] While these unique characteristics of CTs make them an ideal areal unit for studying the role of area-level SEP in environmental health applications, few previous air pollution epidemiological studies have had the ability to apply them.

We leveraged a unique data source of daily emergency department (ED) visits and admissions data in the state of Missouri that included information on patient residential CT to better understand how census tract-level SEP affects the risk of acute CVDM due to ambient PM_2.5_ exposure. We hypothesized that, relative to residents of census tracts with high SEP, residents of census tracts with low SEP will be at an elevated risk for PM_2.5_-related acute CVDM ED visits and admissions.

## Methods

### Emergency Department Visit Data

Patient-level ED visit data were obtained from the Missouri Department of Health and Senior Services for the period 2012-2016. Our definition of an ED visit included visits by patients that were subsequently admitted to the hospital. The study population was restricted to all ED patients who had a residential address geocoded to the CT level in the state of Missouri. Cause-specific ED visits for CVDM were identified as those with primary or secondary International Classification of Diseases (ICD) version 9 codes 390-459 or ICD version 10 codes I00-I99.

Our total sample included 3,539,599 ED visits from 1,393 CTs by Missouri residents during 2012-2016, among which 1,617,158 ED visits were from 641 CTs in urban areas (Figure 1). We defined urban CTs as those contained within counties that comprise Metro-Statistical Areas (MSAs) with at least 1 million residents living in the state of Missouri. These included the St. Louis, MO-IL and Kansas City, MO-KS MSAs. [20]. The Institutional Review Board (IRB) at Emory University approved this study and granted an exemption from informed consent requirements.

**Figure 1.**
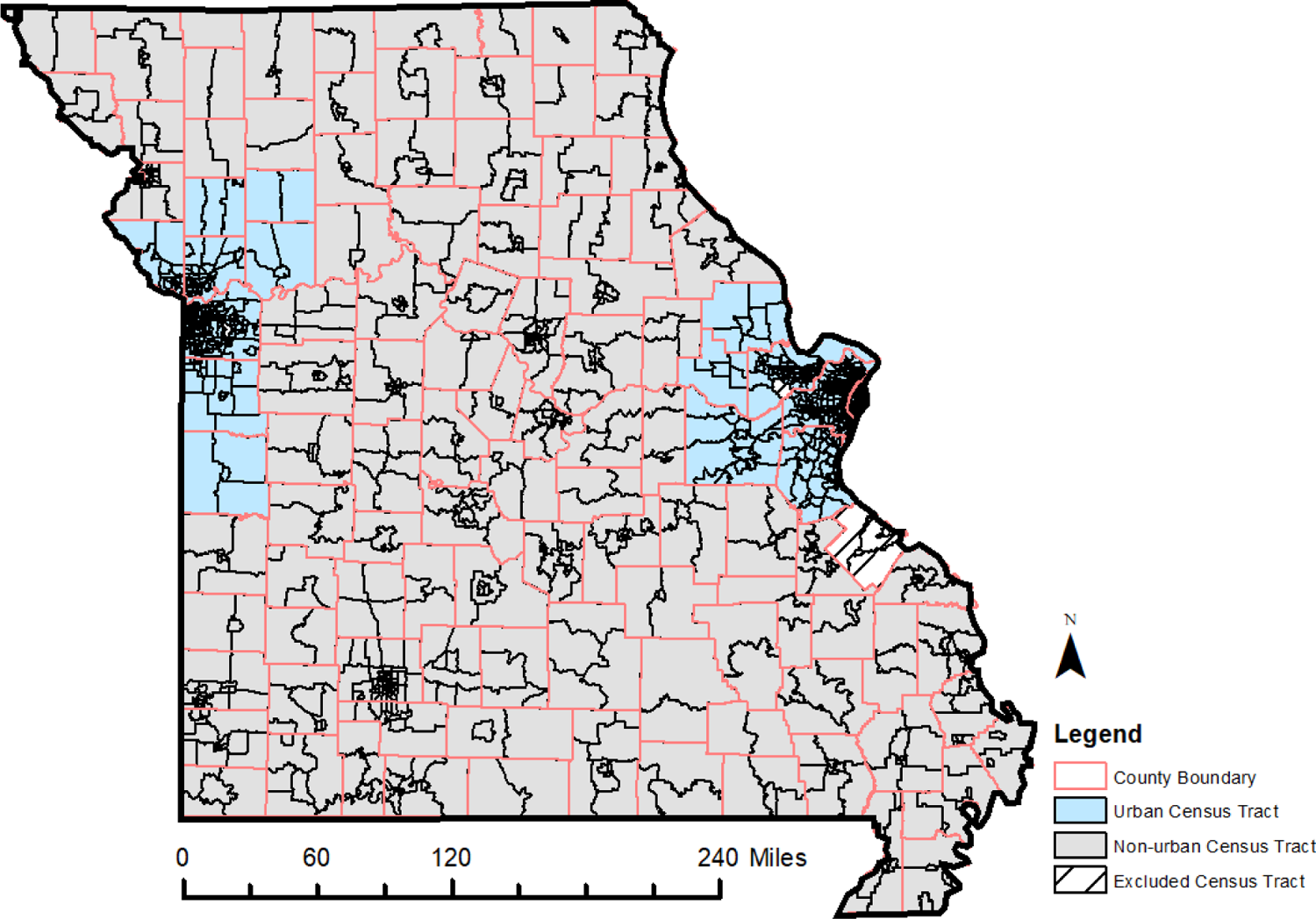
Map of Missouri Counties and Census Tracts Study area for the main analyses. Grey areas represent non-urban census tracts. Blue areas represent urban census tracts. White hash marks represent excluded census tracts (≤50 cardiovascular ED visits over the course of the study or missing socioeconomic data) **A.** Effects of 25-HC on IAV growth kinetics. Ethanol was used as a negative control. *Hmgcr* gene expression and HMGCR protein abundance.

### Ambient PM_2.5_ and Meteorological Data

Daily ambient air pollution data were retrieved from the Socioeconomic Data and Applications Center (SEDAC) for the period 2012 to 2016. The PM_2.5_ product provided daily concentrations at 1km x 1km grid cell resolution, estimated using an ensemble of machine learning algorithms that had good predictive performance (R^2^ > 0.86). [22] Daily CT-level PM_2.5_ concentrations were obtained by averaging the daily concentration of PM_2.5_ from 1km x 1km grid cells with centroids located with each given CT.

Daily 1km x 1km meteorological data were retrieved from the Daily Surface Weather and Climatological Summary (Daymet) [21] and included daily maximum temperature and daily average dewpoint temperature. Census tract-level exposures were estimated by assigning the 1km x 1km Daymet grid cell to the census tract centroid.

### Area-Level Effect Modifiers

Area-level modifiers were obtained from the Census Bureau’s American Fact Finder (AFF) based on the EPA’s EJ Screen tool. [4] For all area level modifiers, we used 2012-2016 5-year estimates at census tracts. We examined five area-level modifiers: (1) the percentage of POC, defined as individuals identifying as not white alone and not white and Hispanic or Latino, (2) the percentage of people for whom poverty status is determined (calculated using yearly Census Bureau poverty thresholds based on family size and number of children [22]), (3) the percentage of people unemployed (among those >16 years of age and in the labor force), (4) the percentage of limited English speaking households (households where no person aged 14 years or older speaks English “very well”), (5) the percentage of people ≥25 years of age without a high school diploma, and (6) the percentage of a county age 65 years and over. Higher percentage values are associated with lower SEP or a larger proportion of elderly CT residents.

### Analytic Methods

We applied a two-stage modeling approach to estimate associations between daily CT-specific PM_2.5_ concentrations and CVD ED visits, as well as to evaluate effect modification by area-level SEP. In Stage 1, associations between pollution concentrations and CVD ED visits were estimated for every census tract in Missouri using conditional logistic regression, matching on year, month, and day of the week of the ED visit. The general structure of each Stage 1 model was:

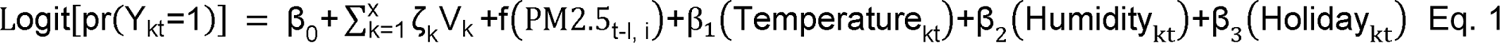

where Y_kt_ indicates whether patient k visited the ED on day t (1 = case; 0 = control day). V_k_ denotes the indicator variables that distinguish the case–control sets for individuals within the study, x is the total number of case–control sets, and ζ_k_ indicates parameters that are specific to the case–control sets (not estimated in conditional logistic regression). We estimated PM_2.5_ effects for same day (lag 0) and 3-day moving average (MA) (lags 0-2) exposures. We chose a 3-day MA of PM_2.5_ as our *a priori* lag structure based on previous work. [23–26] Models included control for maximum temperature and mean dew point using restricted cubic splines with knots at the 25^th^ and 75^th^ percentiles, and holidays using a binary indicator that designated whether or not day t occurred on a holiday.

In Stage 2, we used random effects meta-regression to combine CT-specific PM_2.5_ effect estimates accounting for (1) uncertainty associated with each CT-specific log odds ratio as measured by its asymptotic standard error, and (2) between-CT variability of the true unobserved CT-specific log odds ratio.

We excluded patients with invalid census tract assignments from Stage 1 analysis. Census tracts with Stage 1 models that did not converge because they had fewer than 50 total ED visits during the study period and census tracts with missing SEP data were excluded from Stage 2 analysis (n = 12). After excluding observations with invalid and missing data, the analytic sample contained observations for 3,314,398 (93.6%) individuals from 1,381 CTs (Supplemental Table 1).

In Stage 2, we also explored how area-level social and economic indicators modified the relationship between ambient PM_2.5_ and CVD ED visits by fitting the following meta-regression equation:

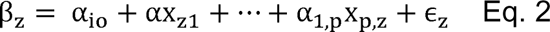

Where β_z_ is the true effect estimate for census tract z (estimated by log odds ratios, β_z_) and X_z,1_… X_z,p_ are area-level modifiers, α_l_, … α_p_ are the corresponding regression coefficients, and ε_z_ represents between-CT variation, assumed to be mean-zero normal. In these models, we used SEP indicators categorized by quartiles. We used the first quartile of each indicator, corresponding to the CTs with highest SEP, as the reference level.

In secondary analyses, we considered analyses restricted to urban counties and warm seasons. We defined the warm season as the five months of the year with the highest average maximum temperature (i.e., May through September). [27] All analyses were done in Stata 17.

## RESULTS

### Descriptive Statistics

Table 1 presents descriptive statistics for PM_2.5_, meteorological data, and area-level SEP for all CTs and urban CTs in Missouri included in the analytic sample. Mean daily ambient PM_2.5_ across all CTs was 8.64 µg/m^3^ and among urban CTs was 8.86 µg/m^3^. PM_2.5_ concentrations were higher during the warm season. Figure 2 displays the distributions of area-level effect modifiers across all CTs and urban CTs, respectively. For both statewide and urban samples, census-tract level SEP data displays a positive skew across all indicators.

**Table 1:**
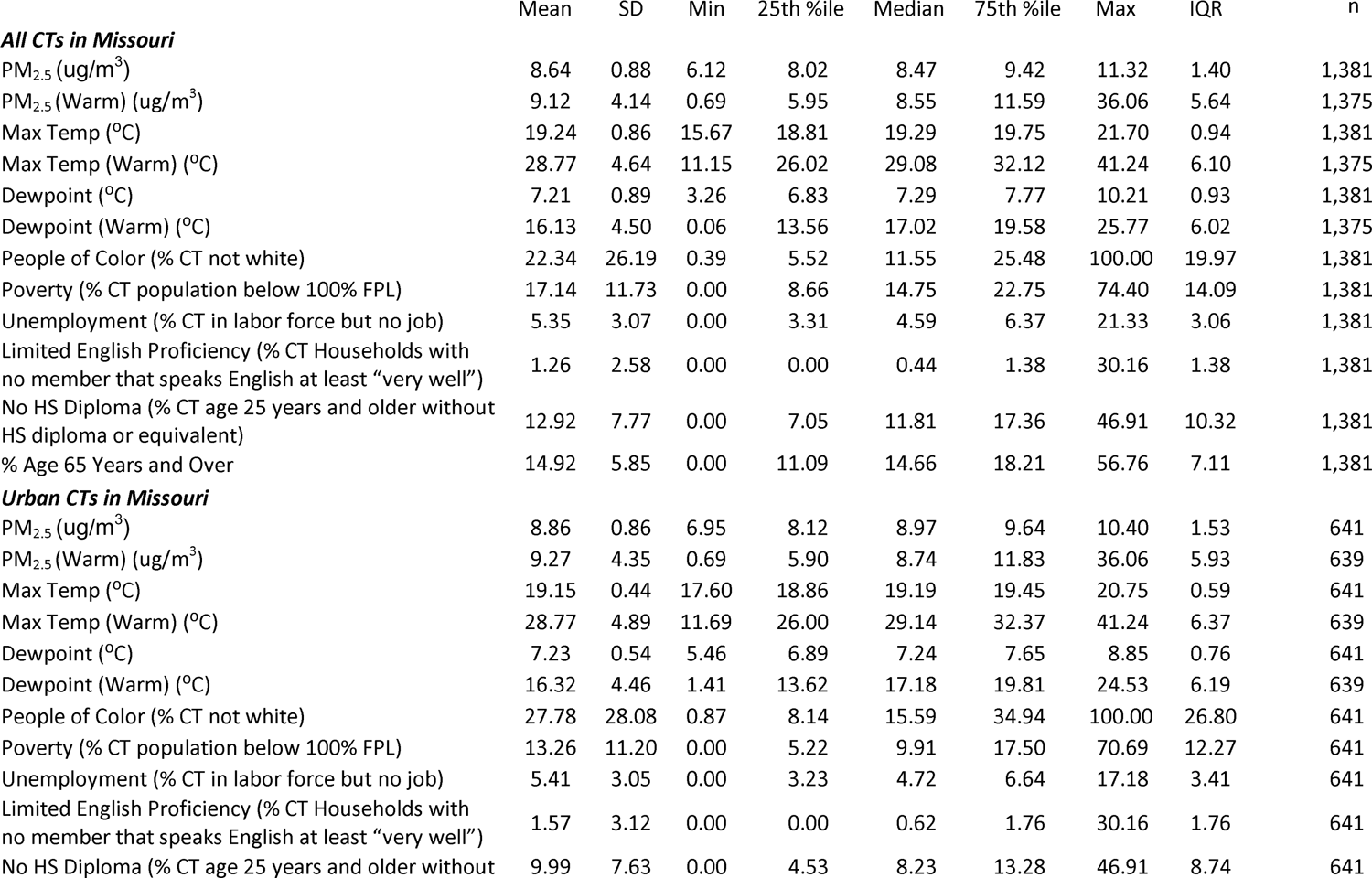
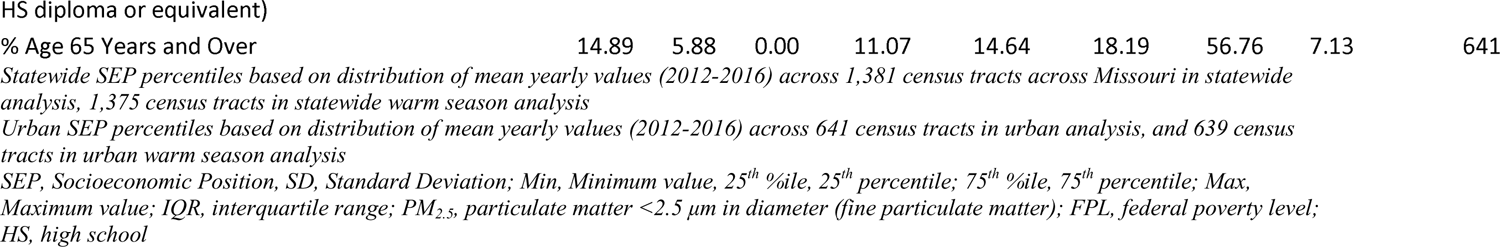
Descriptive Statistics for Census Tract Level PM2.5 Concentrations, Meteorology, and Socioeconomic Position Indicators in Missouri (2012-2016)

**Figure 2.**
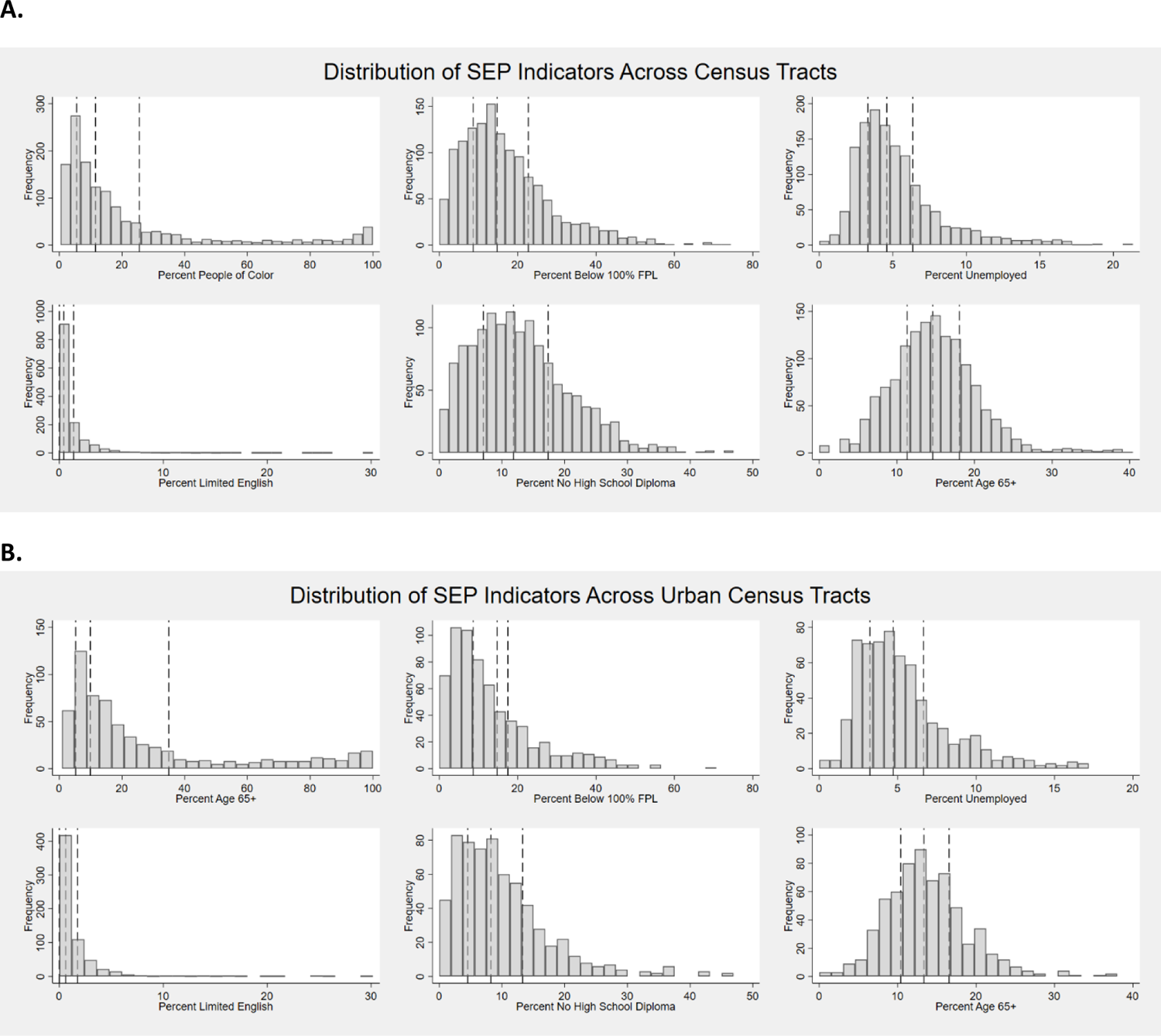
Distribution of Socioeconomic Position Indicators in Missouri (2012-2016) **A.** Represents distributions across the statewide analytic sample (n=1,381 census tracts); **B.** Represents distributions across the statewide analytic sample (n = 641 census tracts); In each histogram the leftmost vertical dashed lines represent the 25^th^ percentile, the central dashed line represents the median, and the rightmost dashed line represents the 75^th^ percentile

Correlations among statewide and urban daily indicators (i.e., PM_2.5_, temperature, dewpoint) can be found in Supplemental Table 2, and correlations between yearly area-level SEP indicators are in Supplemental Table 3. Among SEP indicators, only the percentage of CT residents below 100% of the federal poverty level (FPL) and the percentage of CT residents without a high school diploma (or equivalent) are strongly correlated (r ≥ .70) in both the statewide and urban samples. The percentage of CT residents without a high school diploma (or equivalent) is strongly correlated with the percentage of unemployed CT residents in the statewide sample.

### Overall Associations between PM_2.5_ and ED Visits

Overall, PM_2.5_ was consistently associated with elevated risk of acute CVDM (Table 2), with associations slightly stronger in the warm season than year-round. Specifically, in the warm season, the OR for CVDM ED visits per interquartile range increase (1.40 µg/m^3^) in lag 0 PM_2.5_ was 1.009 (95% CI 1.005, 1.012); the OR based on 3-day MA PM_2.5_ was 1.008, (95% CI 1.004, 1.011). Associations were slightly attenuated when restricted to urban CTs, with a warm season lag 0 OR of 1.007 (95% CI 1.002, 1.011) and 3-day MA OR of 1.005 (95% CI 1.000, 1.010).

**Table 2:**
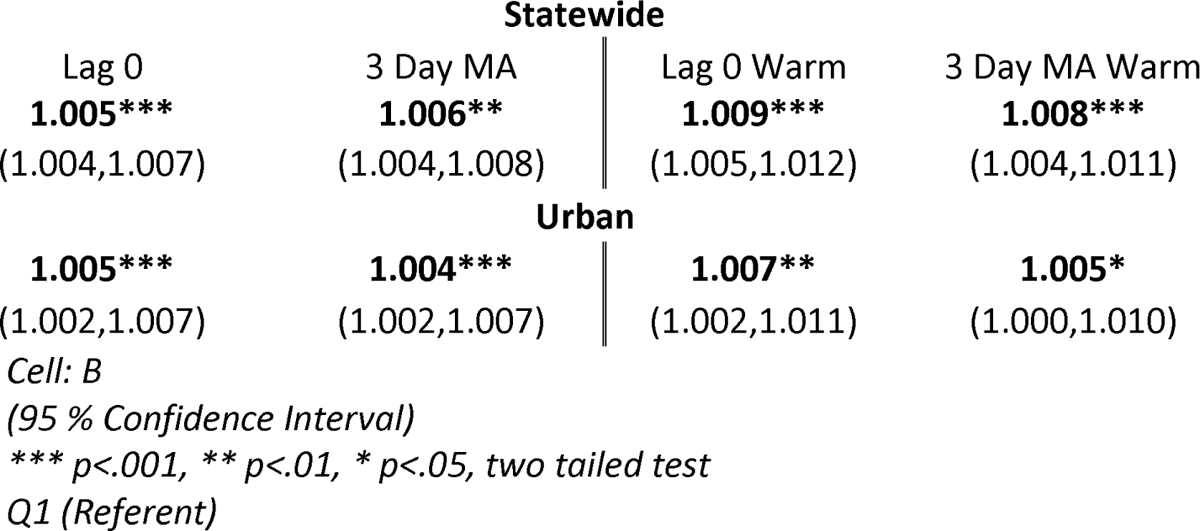
Results for Overall Statewide and Urban Estimates Between Ambient PM_2.5_ Concentration and Cardiovascular ED Visits Across Missouri.

| Statewide |  |  |  |
| --- | --- | --- | --- |
| Lag 0 | 3 Day MA | Lag 0 Warm | 3 Day MA Warm |
| **1.005\*\*\*** | **1.006\*\*** | **1.009\*\*\*** | **1.008\*\*\*** |
| (1.004,1.007) | (1.004,1.008) | (1.005,1.012) | (1.004,1.011) |
| Urban |  |  |  |
| **1.005\*\*\*** | **1.004\*\*\*** | **1.007\*\*** | **1.005\*** |
| (1.002,1.007) | (1.002,1.007) | (1.002,1.011) | (1.000,1.010) |

Cell: B
(95 % Confidence Interval)
\*\*\* $p < .001$ , \*\* $p < .01$ , \* $p < .05$ , two tailed test
Q1 (Referent)

### Effect Modification Analysis

We found evidence of effect modification of PM_2.5_-acute CVDM associations by area-level SEP (statewide results presented in Figure 3 and Supplemental Table 4). In particular, CTs with the lowest levels of poverty had weaker associations [e.g., lag 0 OR of 1.004 (95% CI 1.000, 1.008)] compared to CTs with the highest levels of poverty [e.g., lag 0 OR of 1.010 (95% CI 1.007, 1.014)]. This effect modification was more pronounced during the warm season, with even stronger associations observed for CTs with the highest levels of poverty [e.g., lag 0 OR of 1.015 (95% CI 1.009, 1.022)]. Patterns of stronger associations among CTs of lower SEP were also observed when using SEP indicators for % high school diploma and % unemployed, and again particularly in the warm season.

**Figure 3:**
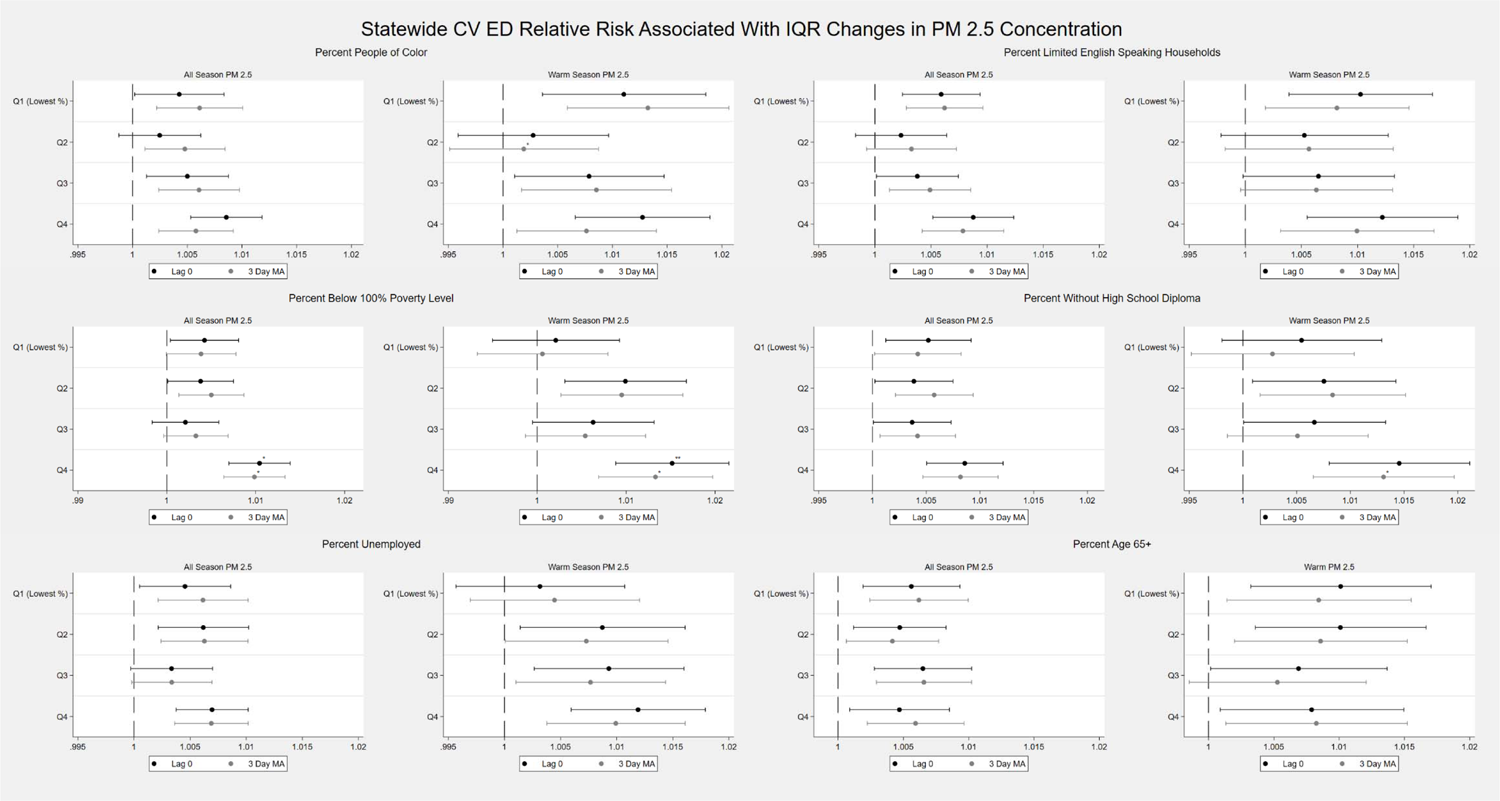
Results for Effect Modification Estimates in Missouri by Socioeconomic Position Indicator and Quartile (n = 1,381) *** p<.001, ** p<.01, * p<.05, two tailed test.

Patterns of effect modification were similar, overall, when restricted to urban CTs (Supplemental Figure 1, Supplemental Table 4). For example, in the urban analysis, the estimated effect of warm season PM_2.5_ on acute CVDM was significantly higher among CTs with the highest levels of poverty [lag 0 OR of 1.015 (95% CI 1.005, 1.026)] compared to CTs with the lowest levels of poverty [lag 0 OR of 0.999 (95% CI 0.992, 1.007)].

In both statewide and urban analyses, we observed some evidence of non-linear effect modification, with weaker PM_2.5_-acute CVDM associations among the middle categories of % POC and % limited English-speaking households than the lowest and highest categories. For example, in the warm season, CTs with % POC in Q2 had the weakest PM_2.5_-acute CVDM association [lag 0 OR of 0.995 (95% CI 0.985, 1.005)] compared to associations for CTs with % POC in Q1, Q3 or Q4 (Supplemental Table 4, Figure 3). Overall, there was little evidence of effect modification by % age 65+ years.

## DISCUSSION AND CONCLUSION

In this analysis of over 3.3 million ED visits in the state of Missouri for cardiovascular conditions, we examined CT-level SEP as a potential modifier for the relationship between ambient PM_2.5_ and CVD ED visits. We used multiple SEP measures to assess potential effect modification due to area-level social factors in analyses considering statewide and urban-only domains.

In overall models we observed that increases in daily PM_2.5_ concentrations significantly increase the odds of visits to the ED for acute CVDM across the state of Missouri in both year-round and warm season-only analyses. Same day (lag 0) effect estimates were similar to 3-day moving average effect estimates, and warm season effect estimates were stronger than their year-round counterparts. Estimated effects in statewide analyses were largely similar to those restricted to urban-only CTs. These findings are consistent with previous literature, which has established that PM_2.5_ is a risk factor for CVDM. [28,29] Previous literature also supports our findings of a seasonal effect of PM_2.5_ on CVDM, but is split on whether cold or warm season exposure presents greater risk. [30–33] Others have suggested that humidity, not temperature, is the primary driver of observed seasonal differences in CVDM associated with PM_2.5_ concentrations. [10,34]

Effect modification varied based on SEP indicator, but results are concurrent with previous literature that suggest CTs may be better suited for understanding how area-level social inequities affect health outcomes. [5,16] The most robust evidence for modification of the PM_2.5_- acute CVDM association in this study was observed when examining the percent of people living below 100% FPL as the SEP indicator. Across all lags and seasons CTs with the highest levels of poverty consistently had an elevated odd of CVDM due to PM_2.5_ compared to CTs with the lowest levels of poverty. Patterns of effect modification were similar when considering percent high school education as the SEP indicator. The pattern of effect modification for some indicators, such as percent POC and percent limited English-speaking households, were not as clear.

The varied nature of results by SEP indicators suggests that not all measures of SEP weigh equally on the risks of PM_2.5_-related CVDM. Rather, SEP reflects access to specific social advantages, and disadvantages, that confer different levels of risk to CT residents. Our finding that poverty is a salient social factor in modifying the association between PM_2.5_ concentrations and CVDM is consistent with other literature, [36] and is seen with other air pollutants. [37,38] The exact mechanisms driving this relationship are unclear, but increased levels of psychosocial stress, reduced access to individual and community resources, or a lack residential green space, all associated with high rates of poverty and low levels of education could be responsible for these findings. [39] Research has also shown that education may be related to adherence to public health guidelines for air quality alerts, which might drive the observed effect modification relationship by education on PM_2.5_-related CVDM in the warm season in this study. [26]

Our finding that, compared to CTs with the lowest and highest percentages of POC, CTs with the second lowest percentages of POC were at lower risk for PM_2.5_-related CVDM during the warm season was unexpected. Although it is difficult to find an exact analogue for this study, previous research has found a similar pattern between air pollution exposure and ED visits when dividing SEP indicators into quartiles. [40] It is important to recognize that the results in these analyses are relative risks, presented by SEP quartile. It is possible that the highest urban SEP quartiles (Q1) have very low overall risks for CVDM associated with PM_2.5_ concentrations during the warm season. A higher relative risk for ED visits in CTs with the fewest POC may reflect a lower overall risk for ED visits relative to CTs with the second lowest rate of POC, as opposed to a true protective effect.

The lack of evidence of the percentage of residents aged 65 years and over functioning as an effect modifier was also unexpected, given the relationship between age, PM_2.5_, exposure and CVDM. [14] In our sample the percentage of CT residents age 65 years and older was inversely correlated with the percentage of CT residents below 100% FPL (Supplemental Table 3). Thus, while age may confer greater risk for individuals, community level risk may be offset by lower levels of poverty and greater access to material resources.

Our study faced several limitations. Our SEP measurements were individual census tract parameters. While we observed some significant effect modification, none of these indicators were able to fully capture the complex nature and interaction of social conditions within CTs. In addition, we only tested six potential area-level SEP modifiers, leaving room for potential omitted variables. Overall risk across different SEP groups may also interfere with the ability to detect effect modification due to area-level SEP. In these analyses we focused on the relationship between PM_2.5_ and CVDM. Although this relationship is important for better understanding acute cardiovascular health, it is possible that other pollutants could be affecting CVDM. Finally, our analysis was limited to the state of Missouri. It is not clear if our results are generalizable to other states. Further, while we included all ED visits for all patients who visited hospitals in Missouri, we cannot be sure of the characteristics of Missouri residents that chose to visit EDs elsewhere.

Our research provides evidence that census tract level poverty rates contribute to vulnerability to PM_2.5_-related CVDM in Missouri, particularly during the warm season. Other SEP indicators, such as rates of high school diploma attainment and unemployment rate, trend in the expected direction but do not achieve significance. Previous literature on the effect modification relationship between SEP and CVDM associated with PM_2.5_ concentrations has been mixed. [14] Our findings suggest that the ability to detect effect modification is sensitive to the SEP measures chosen. Spatial scale may also play an important role in detecting effect modification. Spatial units that are, on average, smaller and align to social and economic boundaries, such as CTs, may be more sensitive to effect modification than those that do not. Moving forward we recommend that researchers incorporate multiple SEP indicators in their research, and test their findings at multiple spatial resolutions.

## Data Availability

Exposure and meteorological data are available online at: https://sedac.ciesin.columbia.edu/data/set/aqdh-pm2-5-concentrations-contiguous-us-1-km-2000-2016 (exposure) https://daymet.ornl.gov/ (meteorological) Patient health information are available upon reasonable request to the author, IRB approval, and State of Missouri Department of Health and Senior Services

## Acknowledgements

The data used in this document/presentation was acquired from the Missouri Department of Health and Senior Services (DHSS). The contents of this document including data analysis, interpretation or conclusions are solely the responsibility of the authors and do not represent the official views of DHSS. We would also like to acknowledge several collaborators. Thank you to Haisu Zhang for help curating exposure and meteorological data, Morgan Lane for her assistance with ensuring compliance with IRB protocols and state-level data use agreements, and Shannon Moss for helping to facilitate access to patient health information.

## Notes

### Competing Interest Statement

The authors have declared no competing interest.

### Funding Statement

NIEHS R01ES027892
NIEHS T32ES012870

### Author Declarations

The Institutional Review Board (IRB) at Emory University approved this study and granted an exemption from informed consent requirements.

## References

1 Chi GC, Hajat A, Bird CE, et al. Individual and Neighborhood Socioeconomic Status and the Association between Air Pollution and Cardiovascular Disease. Environ Health Perspect 2016;124:1840–7. doi:10.1289/EHP199

2 Du Y, Xu X, Chu M, et al. Air particulate matter and cardiovascular disease: the epidemiological, biomedical and clinical evidence. J Thorac Dis 2016;8:E8–19. doi:10.3978/j.issn.2072-1439.2015.11.37

3 Lee B-J, Kim B, Lee K. Air pollution exposure and cardiovascular disease. Toxicol Res 2014;30:71–5. doi:10.5487/TR.2014.30.2.071

4 US EPA. Overview of Socioeconomic Indicators in EJScreen. Published Online First: 21 October 2014.https://www.epa.gov/ejscreen/overview-socioeconomic-indicators-ejscreen (accessed 1 Jun 2023).

5 Bravo MA, Anthopolos R, Bell ML, et al. Racial isolation and exposure to airborne particulate matter and ozone in understudied US populations: Environmental justice applications of downscaled numerical model output. Environ Int 2016;92–93:247–55. doi:10.1016/j.envint.2016.04.008

6 Hicken MT, Payne-Sturges D, McCoy E. Evaluating Race in Air Pollution and Health Research: Race, PM2.5 Air Pollution Exposure, and Mortality as a Case Study. Curr Environ Health Rep 2023;10:1–11. doi:10.1007/s40572-023-00390-y

7 Collins TW, Grineski SE. Racial/ethnic disparities in short-term PM2.5 air pollution exposures in the United States. Environ Health Perspect 2022;130:87701. doi:10.1289/EHP11479

8 Richmond-Bryant J, Mikati I, Benson AF, et al. Disparities in distribution of particulate matter emissions from US coal-fired power plants by race and poverty status after accounting for reductions in operations between 2015 and 2017. Am J Public Health 2020;110:655–61. doi:10.2105/AJPH.2019.305558

9 Kravitz-Wirtz N, Teixeira S, Hajat A, et al. Early-Life Air Pollution Exposure, Neighborhood Poverty, and Childhood Asthma in the United States, 1990–2014. Int J Environ Res Public Health 2018;15:1114. doi:10.3390/ijerph15061114

10 Klompmaker JO, Hart JE, James P, et al. Air pollution and cardiovascular disease hospitalization - Are associations modified by greenness, temperature and humidity? Environ Int 2021;156:106715. doi:10.1016/j.envint.2021.106715

11 Hazlehurst MF, Nurius PS, Hajat A. Individual and Neighborhood Stressors, Air Pollution and Cardiovascular Disease. Int J Environ Res Public Health 2018;15. doi:10.3390/ijerph15030472

12 Bai LI, Shin S, Burnett RT, et al. Exposure to ambient air pollution and the incidence of congestive heart failure and acute myocardial infarction: A population-based study of 5.1 million Canadian adults living in Ontario. Environ Int 2019;132:105004.https://www.sciencedirect.com/science/article/pii/S0160412019310608

13 Jin T, Di Q, Réquia WJ, et al. Associations between long-term air pollution exposure and the incidence of cardiovascular diseases among American older adults. Environ Int 2022;170:107594. doi:10.1016/j.envint.2022.107594

14 Fuller CH, Feeser KR, Sarnat JA, et al. Air pollution, cardiovascular endpoints and susceptibility by stress and material resources: a systematic review of the evidence. Environ Health 2017;16:58. doi:10.1186/s12940-017-0270-0

15 Humphrey JL, Reid CE, Kinnee EJ, et al. Putting Co-Exposures on Equal Footing: An Ecological Analysis of Same-Scale Measures of Air Pollution and Social Factors on Cardiovascular Disease in New York City. Int J Environ Res Public Health 2019;16. doi:10.3390/ijerph16234621

16 Clark LP, Harris MH, Apte JS, et al. National and Intraurban Air Pollution Exposure Disparity Estimates in the United States: Impact of Data-Aggregation Spatial Scale. Environ Sci Technol Lett 2022;9:786–91. doi:10.1021/acs.estlett.2c00403

17 Hicken MT, Adar SD, Hajat A, et al. Air Pollution, Cardiovascular Outcomes, and Social Disadvantage: The Multi-ethnic Study of Atherosclerosis. Epidemiology 2016;27:42–50. doi:10.1097/EDE.0000000000000367

18 Krieger N, Chen JT, Waterman PD, et al. Choosing area based socioeconomic measures to monitor social inequalities in low birth weight and childhood lead poisoning: The Public Health Disparities Geocoding Project (US). J Epidemiol Community Health 2003;57:186–99. doi:10.1136/jech.57.3.186

19 US Census Bureau. Glossary. 2022.https://www.census.gov/programs-surveys/geography/about/glossary.html (accessed 23 Feb 2023).

20 U.S. Census Bureau. Annual Estimates of the Resident Population for Counties in the United States: April 1, 2010 to July 1, 2019”. 2020.

21 NASA. Get Data. Earth DATA. 2021.https://daymet.ornl.gov/getdata (accessed Aug 2022).

22 US Census Bureau. Poverty Thresholds. 2023.https://www.census.gov/data/tables/time-series/demo/income-poverty/historical-poverty-thresholds.html (accessed 18 Jan 2023).

23 O’Lenick CR, Chang HH, Kramer MR, et al. Ozone and childhood respiratory disease in three US cities: evaluation of effect measure modification by neighborhood socioeconomic status using a Bayesian hierarchical approach. Environ Health 2017;16:36. doi:10.1186/s12940-017-0244-2

24 Strickland MJ, Darrow LA, Mulholland JA, et al. Implications of different approaches for characterizing ambient air pollutant concentrations within the urban airshed for time-series studies and health benefits analyses. Environ Health 2011;10:36. doi:10.1186/1476-069X-10-36

25 Strickland MJ, Darrow LA, Klein M, et al. Short-term associations between ambient air pollutants and pediatric asthma emergency department visits. Am J Respir Crit Care Med 2010;182:307–16. doi:10.1164/rccm.200908-1201OC

26 O’Lenick CR, Winquist A, Mulholland JA, et al. Assessment of neighbourhood-level socioeconomic status as a modifier of air pollution-asthma associations among children in Atlanta. J Epidemiol Community Health 2017;71:129–36. doi:10.1136/jech-2015-206530

27 Stowell JD, Sun Y, Spangler KR, et al. Warm-season temperatures and emergency department visits among children with health insurance. Environ Res Health 2023;1:015002. doi:10.1088/2752-5309/ac78fa

28 Basith S, Manavalan B, Shin TH, et al. The Impact of Fine Particulate Matter 2.5 on the Cardiovascular System: A Review of the Invisible Killer. Nanomaterials (Basel) 2022;12. doi:10.3390/nano12152656

29 Lim CC, Hayes RB, Ahn J, et al. Mediterranean Diet and the Association Between Air Pollution and Cardiovascular Disease Mortality Risk. Circulation 2019;139:1766–75. doi:10.1161/CIRCULATIONAHA.118.035742

30 Kioumourtzoglou M-A, Schwartz J, James P, et al. PM2.5 and Mortality in 207 US Cities: Modification by Temperature and City Characteristics. Epidemiology 2016;27:221–7. doi:10.1097/EDE.0000000000000422

31 Talbott EO, Rager JR, Benson S, et al. A case-crossover analysis of the impact of PM2.5 on cardiovascular disease hospitalizations for selected CDC tracking states. Environ Res 2014;134:455–65. doi:10.1016/j.envres.2014.06.018

32 Hsu W-H, Hwang S-A, Kinney PL, et al. Seasonal and temperature modifications of the association between fine particulate air pollution and cardiovascular hospitalization in New York state. Sci Total Environ 2017;578:626–32. doi:10.1016/j.scitotenv.2016.11.008

33 Yitshak-Sade M, James P, Kloog I, et al. Neighborhood Greenness Attenuates the Adverse Effect of PM2.5 on Cardiovascular Mortality in Neighborhoods of Lower Socioeconomic Status. Int J Environ Res Public Health 2019;16. doi:10.3390/ijerph16050814

34 Qiu H, Yu IT-S, Wang X, et al. Cool and dry weather enhances the effects of air pollution on emergency IHD hospital admissions. Int J Cardiol 2013;168:500–5. doi:10.1016/j.ijcard.2012.09.199

35 Danesh Yazdi M, Wang Y, Di Q, et al. Long-Term Association of Air Pollution and Hospital Admissions Among Medicare Participants Using a Doubly Robust Additive Model. Circulation 2021;143:1584–96. doi:10.1161/CIRCULATIONAHA.120.050252

36 Clougherty JE, Humphrey JL, Kinnee EJ, et al. Social Susceptibility to Multiple Air Pollutants in Cardiovascular Disease. Res Rep Health Eff Inst 2021;:1– 71.https://www.ncbi.nlm.nih.gov/pubmed/36004603

37 Adler NE, Newman K. Socioeconomic disparities in health: pathways and policies. Health Aff 2002;21:60–76. doi:10.1377/hlthaff.21.2.60

38 Bernard P, Charafeddine R, Frohlich KL, et al. Health inequalities and place: a theoretical conception of neighbourhood. Soc Sci Med 2007;65:1839–52. doi:10.1016/j.socscimed.2007.05.037

39 Sugerman DE, Keir JM, Dee DL, et al. Emergency health risk communication during the 2007 San Diego wildfires: comprehension, compliance, and recall. J Health Commun 2012;17:698–712. doi:10.1080/10810730.2011.635777

40 Hayes RB, Lim C, Zhang Y, et al. PM2.5 air pollution and cause-specific cardiovascular disease mortality. Int J Epidemiol 2020;49:25–35. doi:10.1093/ije/dyz114

