## Supplemental Information for "An assessment of census-tract level socioeconomic position as a modifier of the relationship between PM 2.5 concentrations and cardiovascular emergency department visits in Missouri"

**Contents**

Supplemental Table 1. ED CV Daily Event Visits in Missouri 2012 – 2016	2

Supplemental Table 2: Spearman Correlation Coefficients for PM 2.5 and Metrological Indicators (Daily Indicators) (2012-2016)	3

Supplemental Table 3: Spearman Correlation Coefficients for SEP Indicators (2012-2016)	4

Supplemental Figure 1. Results for Effect Modification Estimates in Urban Missouri by Socioeconomic Position Indicator and Quartile (n = 641)	5

Supplemental Table 4: Effect Modification Estimates for the Relationship Between Ambient PM_2.5_ Concentration and Cardiovascular ED Visits Across Missouri by Socioeconomic Position Quartile	6

References	9

| Supplemental Table 1. ED CV Daily Event Visits in Missouri 2012 - 2016 | | | | | | | | |
| --- | --- | --- | --- | --- | --- | --- | --- | --- |
| **Statewide** | | | | | | | | |
| Total | | Analytic Sample | | | Descriptive Statistics (Daily Visits) | | | |
| ED Visits | Census Tracts | ED Visits | Census Tracts | % ED in Sample | Mean (SD) | Median | Min | Max |
| 3,539,599 | 1,393 | 3,314,398 | 1,381 | 93.63% | 1844.21 (221.12) | 1,878 | 1,157 | 2,457 |
| **Urban*** | | | | | | | | |
| Total | | Analytic Sample | | | Descriptive Statistics (Daily Visits) | | | |
| ED Visits | Census Tracts | ED Visits | Census Tracts | % ED in Sample | Mean (SD) | Median | Min | Max |
| 1,617,158 | 641 | 1,586,483 | 641 | 98.10% | 883.61 (110.45) | 900 | 544 | 1,198 |

**We defined urban CTs as those contained within counties that comprise Metro-Statistical Areas (MSAs) with at least 1 million residents living in the state of Missouri. These included the St. Louis, MO-IL and Kansas City, MO-KS MSAs.^1^ Because CTs map directly to counties, all CTs included in the urban analysis are entirely contained by the counties that make up the St. Louis and Kansas City MSAs.*

| Supplemental Table 2. Spearman Correlation Coefficients for PM 2.5 and Metrological Indicators (2012-2016) | | | | | | |
| --- | --- | --- | --- | --- | --- | --- |
|  | All Season | | | Warm Season | | |
|  | **Statewide** | | | | | |
|  | PM_2.5_ (μg/m3) | Max Temp (⁰C) | Dewpoint (⁰C) | PM_2.5_ (μg/m3) | Max Temp (⁰C) | Dewpoint (⁰C) |
| PM_2.5_ (μg/m3) | 1.00 |  |  | 1.00 |  |  |
| Max Temp (⁰C) | 0.08 | 1.00 |  | 0.52 | 1.00 |  |
| Dewpoint (⁰C) | 0.31 | 0.87 | 1.00 | 0.34 | 0.67 | 1.00 |
| *n = 1,381* | | | | | | |
|  | **Urban** | | | | | |
|  | PM_2.5_ (μg/m3) | Max Temp (⁰C) | Dewpoint (⁰C) | PM_2.5_ (μg/m3) | Max Temp (⁰C) | Dewpoint (⁰C) |
| PM_2.5_ (μg/m3) | 1.00 |  |  | 1.00 |  |  |
| Max Temp (⁰C) | 0.47 | 1.00 |  | 0.64 | 1.00 |  |
| Dewpoint (⁰C) | 0.34 | 0.85 | 1.00 | 0.16 | 0.61 | 1.00 |
| *n = 641* | | | | | | |
| *PM_2.5,_ Particulate matter ≤ 2.5 micrometers in diameter*  *Data is collected or modeled daily* | | | | | | |

| Supplemental Table 3: Spearman Correlation Coefficients for SEP Indicators (2012-2016) | | | | | | |
| --- | --- | --- | --- | --- | --- | --- |
| **Statewide** | | | | | | |
|  | POC | Poverty | Unemployment | English Prof. | Education | Age 65+ |
| POC | 1.00 |  |  |  |  |  |
| Poverty | 0.53 | 1.00 |  |  |  |  |
| Unemployment | 0.42 | 0.72 | 1.00 |  |  |  |
| English Prof. | 0.34 | 0.19 | 0.06 | 1.00 |  |  |
| Education | 0.29 | 0.78 | 0.71 | 0.10 | 1.00 |  |
| Age 65+ | -0.21 | -0.15 | -0.21 | -0.01 | -0.07 | 1.00 |
| *n = 1,381* | | | | | | |
| **Urban** | | | | | | |
|  | POC | Poverty | Unemployment | English Prof. | Education | Age 65+ |
| POC | 1.00 |  |  |  |  |  |
| Poverty | 0.32 | 1.00 |  |  |  |  |
| Unemployment | 0.36 | 0.57 | 1.00 |  |  |  |
| English Prof. | 0.39 | 0.11 | 0.07 | 1.00 |  |  |
| Education | 0.07 | 0.74 | 0.50 | -0.03 | 1.00 |  |
| Age 65+ | -0.43 | -0.10 | -0.23 | -0.19 | 0.06 | 1.00 |
| *n = 641* | | | | | | |
| *Data is collected/estimated yearly*  *POC, percent people of color; percent below 100% federal poverty level, English Prof., percent limited English proficiency; Education, percent ≥ 25 years without high school diploma or equivalent; Age 65+, percent ≥ age 65* | | | | | | |

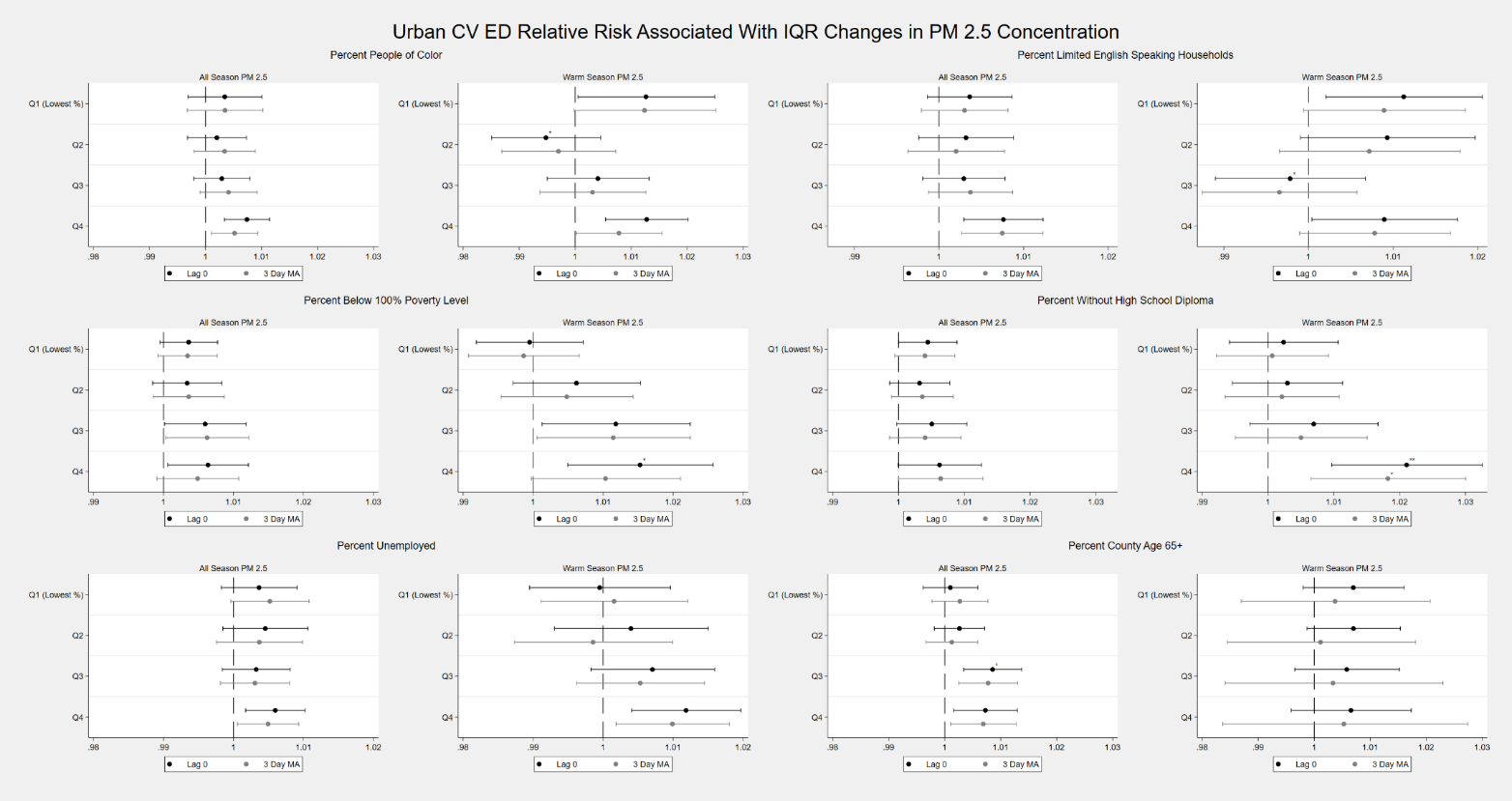

*Supplemental Figure 1. Results for Effect Modification Estimates in Urban Missouri by Socioeconomic Position Indicator and Quartile (n = 641)*

**** p<.001, ** p<.01, * p<.05, two tailed test*

*Q1 (Referent)*

| Supplemental Table 4: Effect Modification Estimates for the Relationship Between Ambient PM_2.5_ Concentration and Cardiovascular ED Visits Across Missouri by Socioeconomic Position Quartile | | | | | | | | |
| --- | --- | --- | --- | --- | --- | --- | --- | --- |
|  | Urban Census Tracts | | | | Statewide | | | |
|  | ***Census Tract Percent People of Color*** | | | | | | | |
|  | Lag 0 | 3 Day MA | Lag 0 Warm | 3 Day MA Warm | Lag 0 | 3 Day MA | Lag 0 Warm | 3 Day MA Warm |
| Q 1 (Lowest %) | 1.003 | 1.003 | 1.013 | 1.001 | 1.005 | 1.004 | 1.011 | 1.013 |
|  | (0.997,1.010) | (0.997,1.010) | (1.001,1.025) | (0.992,1.009) | (1.001,1.009) | (1.000,1.008) | (1.004,1.019) | (1.006,1.021) |
| Q 2 | 1.002 | 1.003 | **0.995*** | 1.002 | 1.004 | 1.006 | 1.003 | **1.002*** |
|  | (0.997,1.007) | (0.998,1.009) | (0.985,1.005) | (0.993,1.011) | (1.000,1.007) | (1.002,1.009) | (0.996,1.010) | (0.995,1.009) |
| Q 3 | 1.003 | 1.004 | 1.004 | 1.005 | 1.004 | 1.004 | 1.008 | 1.009 |
|  | (0.998,1.008) | (0.999,1.009) | (0.995,1.013) | (0.995,1.015) | (1.000,1.007) | (1.001,1.008) | (1.001,1.015) | (1.002,1.015) |
| Q 4 | 1.007 | 1.005 | 1.013 | 1.018 | 1.009 | 1.008 | 1.013 | 1.008 |
|  | (1.003,1.011) | (1.001,1.009) | (1.005,1.020) | (1.006,1.030) | (1.005,1.012) | (1.005,1.012) | (1.007,1.019) | (1.001,1.014) |
|  | ***Census Tract Percent Below 100% Federal Poverty Level*** | | | | | | | |
|  | Lag 0 | 3 Day MA | Lag 0 Warm | 3 Day MA Warm | Lag 0 | 3 Day MA | Lag 0 Warm | 3 Day MA Warm |
| Q 1 (Lowest %) | 1.004 | 1.003 | 0.999 | 0.999 | 1.004 | 1.004 | 1.002 | 1.001 |
|  | (1.000,1.008) | (0.999,1.008) | (0.992,1.007) | (0.991,1.007) | (1.000,1.008) | (1.000,1.008) | (0.995,1.009) | (0.993,1.008) |
| Q 2 | 1.003 | 1.004 | 1.006 | 1.005 | 1.004 | 1.005 | 1.01 | 1.009 |
|  | (0.998,1.008) | (0.999,1.009) | (0.997,1.015) | (0.995,1.014) | (1.000,1.008) | (1.001,1.009) | (1.003,1.017) | (1.003,1.016) |
| Q 3 | 1.006 | 1.006 | 1.012 | 1.011 | 1.002 | 1.003 | 1.006 | 1.005 |
|  | (1.000,1.012) | (1.000,1.012) | (1.001,1.022) | (1.001,1.022) | (0.998,1.006) | (1.000,1.007) | (0.999,1.013) | (0.999,1.012) |
| Q 4 | 1.006 | 1.005 | **1.015*** | 1.010 | **1.010*** | **1.010*** | **1.015**** | **1.013*** |
|  | (1.001,1.012) | (0.999,1.011) | (1.005,1.026) | (1.000,1.021) | (1.007,1.014) | (1.006,1.013) | (1.009,1.022) | (1.007,1.020) |
|  | ***Census Tract Percent Unemployed (> 16 years, in labor force)*** | | | | | | | |
|  | Lag 0 | 3 Day MA | Lag 0 Warm | 3 Day MA Warm | Lag 0 | 3 Day MA | Lag 0 Warm | 3 Day MA Warm |
| Q 1 (Lowest %) | 1.004 | 1.005 | 1.000 | 1.002 | 1.005 | 1.006 | 1.003 | 1.004 |
|  | (0.998,1.009) | (1.000,1.011) | (0.989,1.010) | (0.991,1.012) | (1.000,1.009) | (1.002,1.010) | (0.996,1.011) | (0.997,1.012) |
| Q 2 | 1.005 | 1.004 | 1.004 | 0.999 | 1.006 | 1.006 | 1.009 | 1.007 |
|  | (0.998,1.011) | (0.998,1.010) | (0.993,1.015) | (0.987,1.010) | (1.002,1.010) | (1.002,1.010) | (1.001,1.016) | (1.000,1.015) |
| Q 3 | 1.003 | 1.003 | 1.007 | 1.005 | 1.003 | 1.003 | 1.009 | 1.008 |
|  | (0.998,1.008) | (0.998,1.008) | (0.998,1.016) | (0.996,1.014) | (1.000,1.007) | (1.000,1.007) | (1.003,1.016) | (1.001,1.014) |
| Q 4 | 1.006 | 1.005 | 1.012 | 1.010 | 1.007 | 1.007 | 1.012 | 1.010 |
|  | (1.002,1.010) | (1.001,1.009) | (1.004,1.020) | (1.002,1.018) | (1.004,1.010) | (1.004,1.010) | (1.006,1.018) | (1.004,1.016) |
|  | ***Census Tract Percent Limited English-Speaking Households*** | | | | | | | |
|  | Lag 0 | 3 Day MA | Lag 0 Warm | 3 Day MA Warm | Lag 0 | 3 Day MA | Lag 0 Warm | 3 Day MA Warm |
| Q 1 (Lowest %) | 1.004 | 1.003 | 1.011 | 1.009 | 1.006 | 1.006 | 1.01 | 1.008 |
|  | (0.999,1.009) | (0.998,1.008) | (1.002,1.021) | (0.999,1.019) | (1.002,1.009) | (1.003,1.010) | (1.004,1.017) | (1.002,1.015) |
| Q 2 | 1.003 | 1.002 | 1.009 | 1.007 | 1.002 | 1.003 | 1.005 | 1.006 |
|  | (0.998,1.009) | (0.996,1.008) | (0.999,1.020) | (0.997,1.018) | (0.998,1.006) | (0.999,1.007) | (0.998,1.013) | (0.998,1.013) |
| Q 3 | 1.003 | 1.004 | **0.998*** | 0.997 | 1.004 | 1.005 | 1.007 | 1.006 |
|  | (0.998,1.008) | (0.999,1.009) | (0.989,1.007) | (0.987,1.006) | (1.000,1.007) | (1.001,1.009) | (1.000,1.013) | (1.000,1.013) |
| Q 4 | 1.008 | 1.007 | 1.009 | 1.008 | 1.009 | 1.008 | 1.012 | 1.010 |
|  | (1.003,1.012) | (1.003,1.012) | (1.000,1.018) | (0.999,1.017) | (1.005,1.012) | (1.004,1.011) | (1.006,1.019) | (1.003,1.017) |
|  | ***Census Tract Percent People Age 25 Years and Older Without a High School Diploma*** | | | | | | | |
|  | Lag 0 | 3 Day MA | Lag 0 Warm | 3 Day MA Warm | Lag 0 | 3 Day MA | Lag 0 Warm | 3 Day MA Warm |
| Q 1 (Lowest %) | 1.004 | 1.004 | 1.002 | 1.001 | 1.005 | 1.004 | 1.005 | 1.003 |
|  | (1.000,1.009) | (0.999,1.009) | (0.994,1.011) | (0.992,1.009) | (1.001,1.009) | (1.000,1.008) | (0.998,1.013) | (0.995,1.010) |
| Q 2 | 1.003 | 1.004 | 1.003 | 1.002 | 1.004 | 1.006 | 1.008 | 1.008 |
|  | (0.999,1.008) | (0.999,1.008) | (0.995,1.011) | (0.993,1.011) | (1.000,1.007) | (1.002,1.009) | (1.001,1.014) | (1.002,1.015) |
| Q 3 | 1.005 | 1.004 | 1.007 | 1.005 | 1.004 | 1.004 | 1.007 | 1.005 |
|  | (1.000,1.010) | (0.999,1.009) | (0.997,1.017) | (0.995,1.015) | (1.000,1.007) | (1.001,1.008) | (1.000,1.013) | (0.999,1.012) |
| Q 4 | 1.006 | 1.006 | **1.021**** | **1.018*** | 1.009 | 1.008 | 1.015 | **1.013*** |
|  | (1.000,1.013) | (1.000,1.013) | (1.010,1.033) | (1.006,1.030) | (1.005,1.012) | (1.005,1.012) | (1.008,1.021) | (1.007,1.020) |
|  | ***Census Tract Percent Aged 65 Years and Older*** | | | | | | | |
|  | Lag 0 | 3 Day MA | Lag 0 Warm | 3 Day MA Warm | Lag 0 | 3 Day MA | Lag 0 Warm | 3 Day MA Warm |
| Q 1 (Lowest %) | 1.001 | 1.003 | 1.007 | 1.004 | 1.006 | 1.006 | 1.01 | 1.008 |
|  | (0.996,1.006) | (0.998,1.008) | (0.998,1.016) | (0.987,1.021) | (1.002,1.009) | (1.002,1.010) | (1.003,1.017) | (1.001,1.016) |
| Q 2 | 1.003 | 1.001 | 1.007 | 1.001 | 1.005 | 1.004 | 1.01 | 1.009 |
|  | (0.998,1.007) | (0.997,1.006) | (0.999,1.015) | (0.984,1.018) | (1.001,1.008) | (1.001,1.008) | (1.004,1.017) | (1.002,1.015) |
| Q 3 | **1.009*** | 1.008 | 1.006 | 1.003 | 1.006 | 1.007 | 1.007 | 1.005 |
|  | (1.003,1.014) | (1.003,1.013) | (0.996,1.015) | (0.984,1.023) | (1.003,1.010) | (1.003,1.010) | (1.000,1.014) | (0.999,1.012) |
| Q 4 | 1.007 | 1.007 | 1.007 | 1.005 | 1.005 | 1.006 | 1.008 | 1.008 |
|  | (1.002,1.013) | (1.001,1.013) | (0.996,1.017) | (0.984,1.027) | (1.001,1.009) | (1.002,1.010) | (1.001,1.015) | (1.001,1.015) |
| Observations  (Census Tracts) | 641 | | 639 | | 1381 | | 1375 | |
| *Cell: Β (95 % Confidence Interval)*  **** p<.001, ** p<.01, * p<.05, two tailed test*  *Q1 (Referent)* | | | | | | | | |

1. U.S. Census Bureau. Annual Estimates of the Resident Population for Counties in the United States: April 1, 2010 to July 1, 2019". 2020.
